# Hide and Seek: Privacy-Preserving Artificial Intelligence with a Feasibility Study in Rare Disease Diagnosis

**DOI:** 10.64898/2026.01.15.26344228

**Authors:** Sivaraman Rajaganapathy, Jennifer St. Sauver, Filippo Pinto e Vairo, Prasad G. Iyer, Hongfang Liu, Jungwei W. Fan

**Affiliations:** Department of Artificial Intelligence and Informatics, Mayo Clinic; Division of Epidemiology, Department of Quantitative Health Sciences, Mayo Clinic; Center for Clinical and Translational Science, Mayo Clinic; Division of Gastroenterology and Hepatology, Department of Medicine, Mayo Clinic; Department of Health Data Science and Artificial Intelligence, University of Texas Health Science Center at Houston

**Keywords:** Patient Data Privacy, Data Encryption, Artificial Intelligence, Rare Diseases, Large Language Models, Information Storage and Retrieval

## Abstract

**Background:** Integrating advanced artificial intelligence (AI) into clinical decision-support often requires the sharing of sensitive patient data with external services, raising privacy concerns. Homomorphic encryption (HE) allows computing directly on encrypted data, without revealing the underlying patient information.

**Objectives:** To develop a large language model (LLM)-assisted diagnosis framework while preserving patient privacy in the clinical text analysis, by leveraging HE and using rare disease (RD) diagnosis as a representative application. To demonstrate HE does not hinder the system performance.

**Materials and Methods:** Texts from patient histories and a RD knowledge base were embedded by LLMs into vectors, then encrypted using HE to obscure private information while retaining the semantic nuances. Diagnostic recommendations were generated by computing and ranking the similarities between the patient history and RD vectors in the encrypted space. The system was evaluated using 50 synthetic case reports (5 RDs, each with 10 reports).

**Results:** Applying HE did protect private information from reverse-embedding attacks. HE imposed little disruption to the diagnostic accuracy, with normalized discounted cumulative gains (nDCG) of 0.6108 ± 0.3412 (encrypted) versus 0.6083 ± 0.3415 (unencrypted). The accuracy and computational performance were tunable and consistent, as demonstrated across five different LLMs.

**Discussion:** Our privacy-preserving framework opens tremendous opportunities toward hosting and serving powerful AI solutions across institution boundaries, which would remove the need for local deidentification and incentivize users to access secure external decision-support services.

**Conclusions:** Integrating HE with LLM retrieval can promote the dissemination of nonredundant, high-capacity AI services by preserving both privacy and accuracy.

## 1. Introduction

Recent developments in large language models (LLMs) and broadly artificial intelligence (AI) have advanced the capacity of decision-support solutions. Seamless integration of LLM into clinical decision-support often requires access to protected health information (PHI) and raises critical security concerns. Sending PHI to outside entities for AI solutions would risk data breaches, which have more than tripled (216 to 745) from 2010 to 2023 in the United States (US) healthcare sector [1]. Countermeasures that are commonly adopted include sticking with on-premises AI deployment, especially for small to medium sized organizations that cannot afford a private cloud service and the associated costs. However, the on-prem approach untowardly limits the options in terms of leveraging more powerful, specialized AI tools while reinforcing siloed, suboptimal efforts at a local scale. Taking rare diseases (RDs) as an example: Despite the collective sum of 400 million people affected by RDs worldwide and a hefty economic burden of $997 billion in the US [2], the individual RDs are characterized by very low prevalence and subjective to an average diagnostic delay of approximately five years [3]. A major barrier to diagnosis is that many patients and primary care providers do not have an easily accessible yet private mechanism to share PHI for consultation, along with other challenges such as the inherent complexity of RDs, heterogeneous clinical presentations, and limited clinician familiarity [4,5]. This would not be the case if there had been centrally hosted RD diagnosis services that operate on the best technology and knowledge while being “blind to” any private information sent from the users.

To achieve the said privacy-preserving AI for clinical text, solutions have been proposed and deidentification (DeID) [6,7] is of top relevance in this context. While DeID by modern AI has achieved high accuracy, it does not guarantee 100% clearance of PHI and can corrupt the useful contents upon aggressive scrubbing. To seek a balanced solution for both strict privacy and minimum burden to the users, we revisited existing techniques and recognized that encryption not only fits well in the current AI stack, where most information is vectorized (as embeddings), but provides some of the strongest formal notions of privacy [6]. Specifically, homomorphic encryption (HE) enables mathematical operations directly on encrypted embeddings and easier AI integration in comparison to conventional encryption schemes. Therefore, we propose combining HE and LLMs to develop a retrieval framework that performs similarity searches using embeddings entirely in the encrypted space. Sending secure queries to AI-mediated knowledge bases (e.g., diagnosis guidelines) allows computation-intensive jobs to be outsourced to third-party decision-support services without revealing sensitive information. Such a framework can be very useful to the Clinical and Translational Science Awards (CTSA) network, for the privacy-preserving feature would encourage developing, hosting, and sharing AI services unique to institution strengths and resources. Centralizing such efforts should reduce redundancy and waste, while allowing users to benefit from more mature solutions in specialized areas.

This study demonstrated the integration of HE and LLM-based diagnostic reasoning to be feasible, especially that the retrieval performance was maintained under encryption. Our use case focused on RD diagnosis from patient history in clinical notes, where privacy-preserving was essential. We further validated that our framework worked for different LLMs and that all computations were completed within practical time and computational resources.

## 2. Materials and Methods

### 2.1. Overview

The proposed framework for privacy-preserving diagnostic retrieval is shown in **Figure 1**, where the client-side processes are depicted at left and the service side at the right. At the client side, a patient’s clinical note is first converted to an LLM embedding vector, referred to as the *query vector*. The client also generates a unique pair of keys for the encryption – the private key is retained by the client, and the public which is used to encrypt the query vector and shared with the server. On the other end, the server uses an LLM to convert an entire disease reference knowledge base into a vector database. In response to a query from the client, along with the public key, the server transforms the vector database into a compatible encrypted space. The server then computes the cosine similarities between the query vector and the vector database entries directly in the encrypted space. Lastly, the encrypted similarity values are returned and decrypted by the client using the private key, followed by ranking the top hit disease entries to complete the diagnostic retrieval. Note that the exact text and its real meaning from the client are never revealed to the server in this framework.

**Figure 1:**
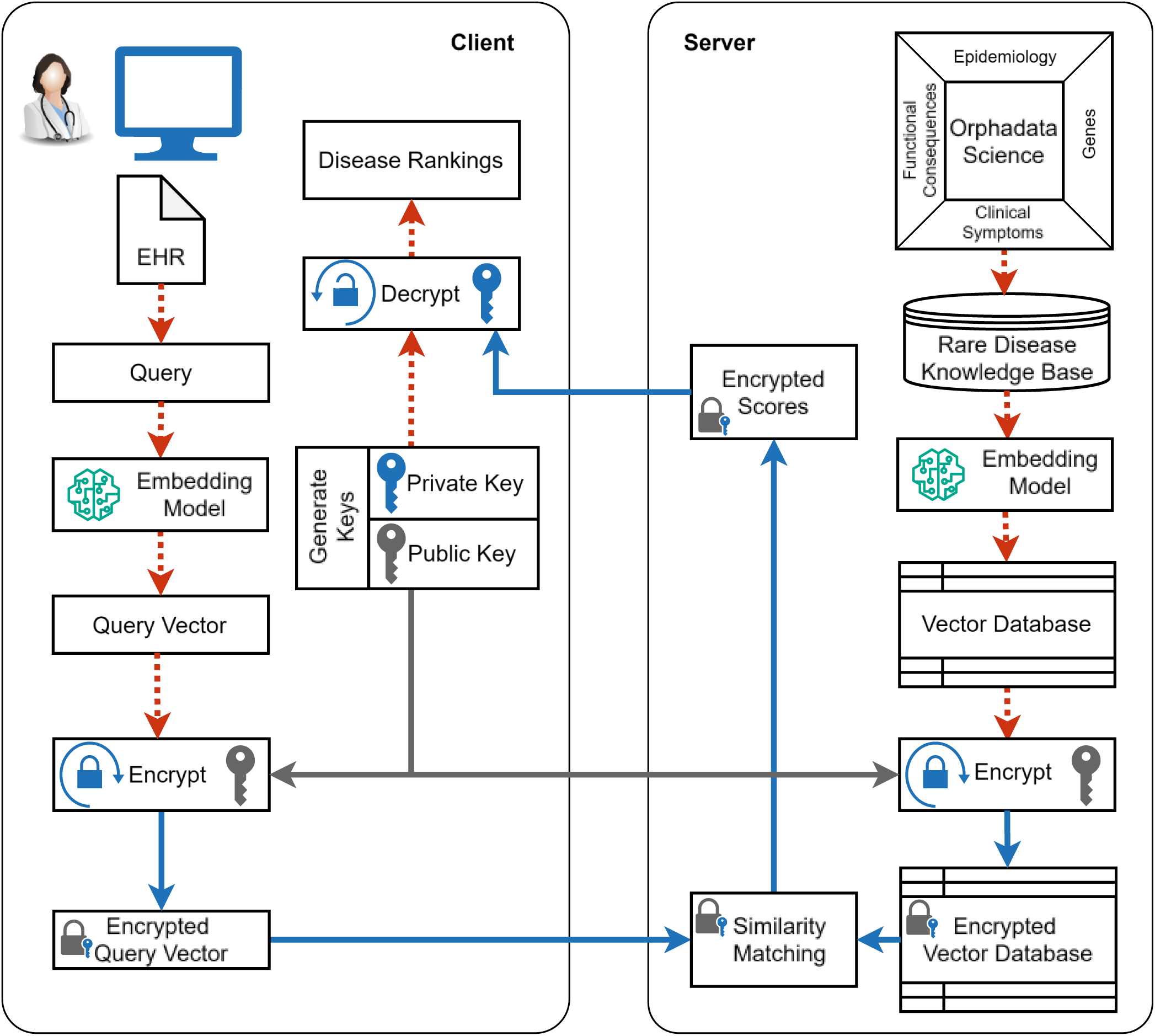
Privacy preserving retrieval with homomorphic encryption (HE). The process comprises steps performed on client and server sides. Here we illustrate the framework through a rare disease diagnosis application, with a custom disease vector database created from the Orphadata Science database. The red dashed arrows represent unencrypted data and the blue solid arrows represent secure encrypted data, as elaborated in the following. A query from the client, which may include a whole clinical note with private information, is transformed into a query vector using an embedding model. The client subsequently generates a unique pair of encryption keys: a public key that is used to encrypt the query vector and shared with the server; a private key that is kept secure later for decrypting the server response back to human-readable information. The server uses the client’s public key to encrypt its vector database and then computes the similarity scores of the query against the vector database in the encrypted space. The encrypted similarity scores are returned to the client, which can decrypt the scores to obtain the retrieval rankings.

### 2.2. Rare disease knowledge base

We used the *Orphadata Science* [8,9], a publicly accessible database providing structured, expert-curated information from the *Orphanet Rare Disease Knowledge Base*. The database contains diverse information associated with over 6,172 unique RDs [9]. We created RD profiles by linking each unique disease with a summary text comprising the associated genes and clinical symptoms. Subsequently, an LLM is used to create an embedding vector from the summary text for each of the RDs, together making our final RD vector database. We used the Python library *Sentence-Transformer* to create the RD vector database [10]. Since the library allows plugging in a range of different LLMs, we experimented with several models of varying sizes (80 MB to 15 GB) and embedding dimensions (384 to 4096). A brief overview of the models is provided in **SM Table 1**.

**Table 1:**
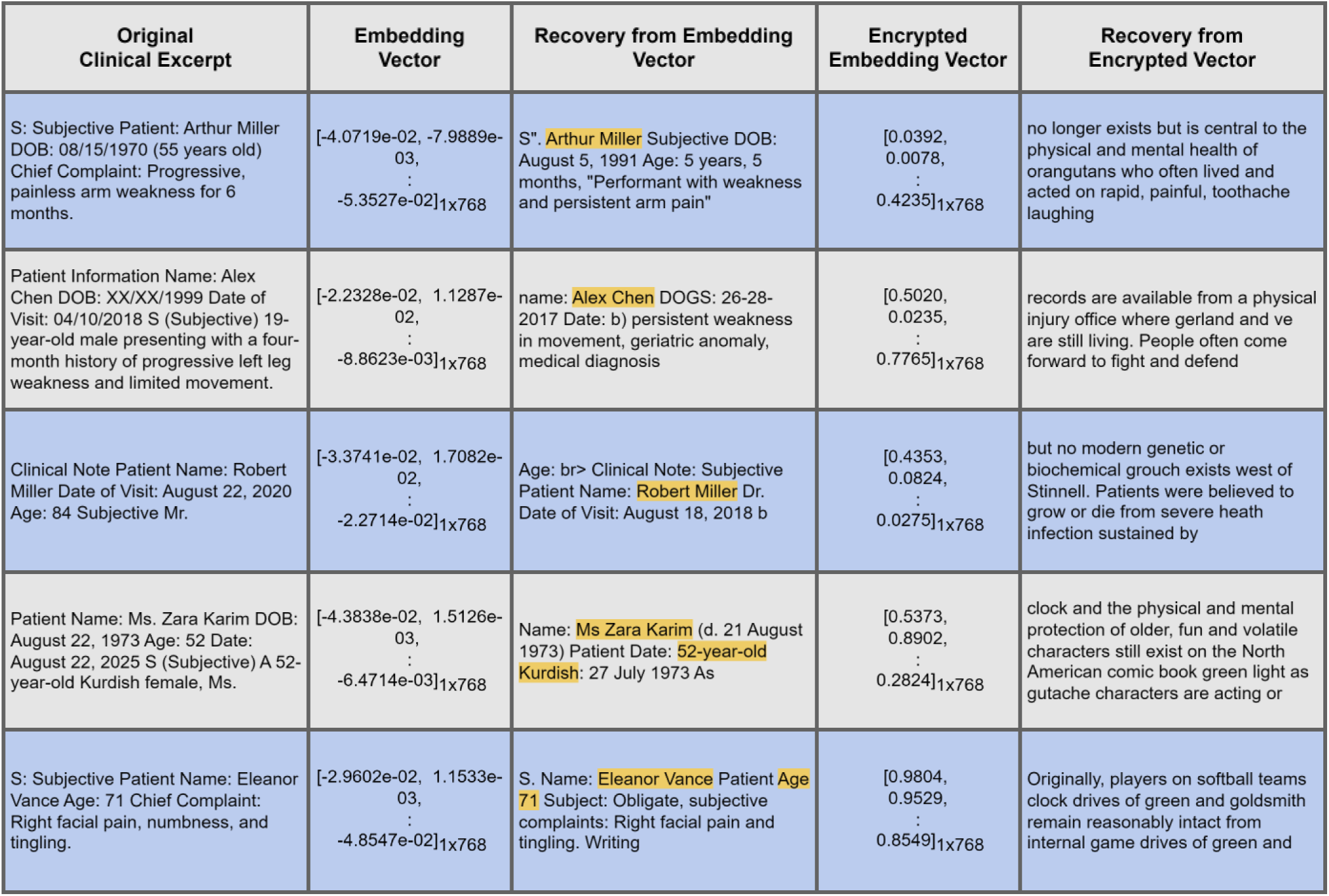
Vulnerability and prevention of privacy leakage from embedding vectors. Excerpts of synthetic clinical notes, corresponding embedding vectors, and the recovered text are shown in the first three columns respectively. Recovered private information is highlighted in yellow. In the fourth and fifth columns, the encrypted embedding vector and the results of a recovery attempt are shown, wherein no meaningful information is revealed.

### 2.3. Rare disease test cases

To demonstrate and validate of our framework, we created synthetic clinical notes as input to simulate a privacy-preserving RD diagnosis application. Based on the authors’ own research interest, five RDs were chosen for the experiments: amyotrophic lateral sclerosis (ALS), fibrodysplasia ossificans progressiva (FOP), idiopathic pulmonary fibrosis (IPF), primary sclerosing cholangitis (PSC), and trigeminal neuralgia (TN). Ten synthetic notes were generated for each of the five RDs (process shown in **Figure 2**) to serve as queries for the diagnostic retrieval. We used the public version of the LLM, *Gemini 2.5 Flash,* specifically prompted to follow the Subjective, Objective, Assessment, Plan (SOAP) structure [11]. As guiding blueprints of realistic medical histories, we supplied *Gemini 2.5 Flash* with case presentation sections selected from full-text PubMed case reports of the five RDs. We ensured that no text revealing the final diagnosis is present in the synthetic notes, so as to simulate the inputs of undiagnosed cases in the task. Accordingly, we extracted only the Subjective and Objective sections of the LLM-synthesized clinical notes for testing the diagnostic retrieval framework. In addition, we prompted the LLM to inject mock patient name in each synthetic note, since the PubMed-sourced case reports did not contain any PHI. Note that the encryption can protect any PHI in the text, and using patient name is just for demonstration.

**Figure 2:**
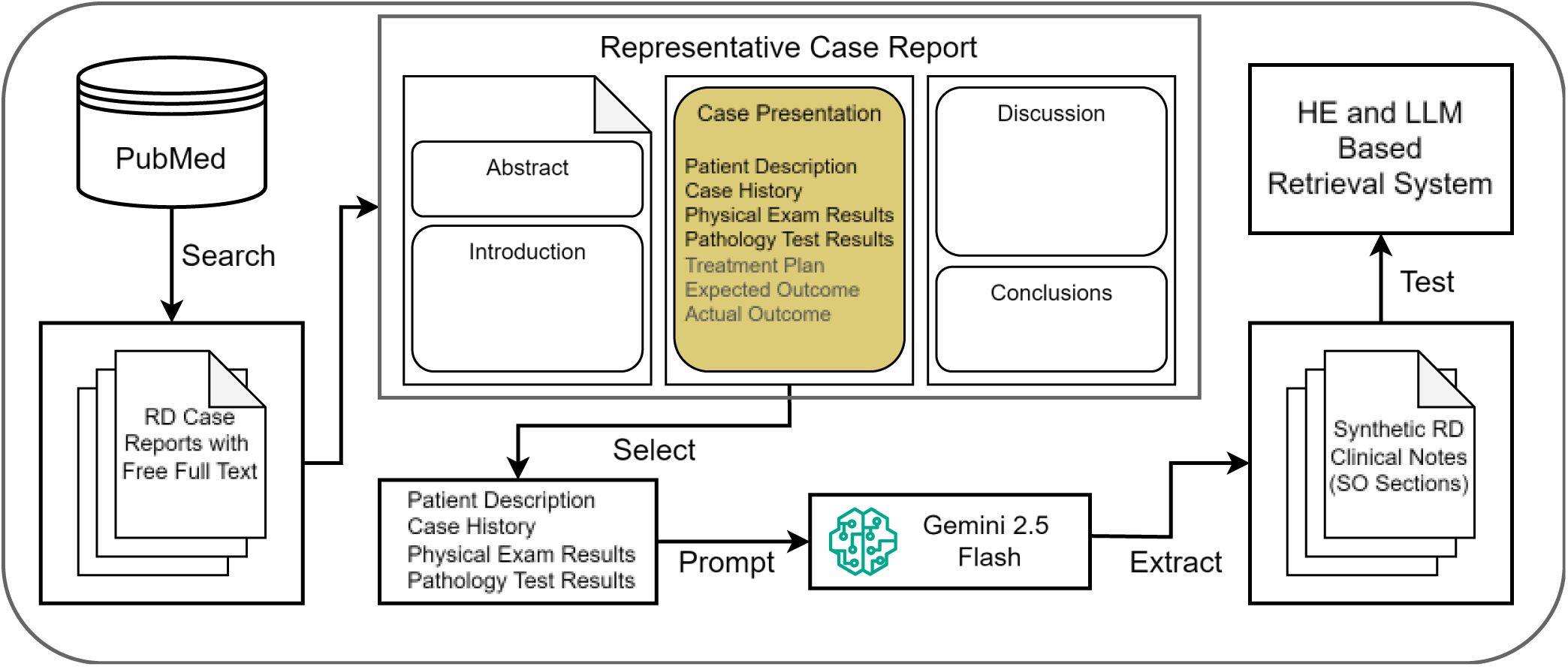
Generation of synthetic test clinical notes. The flowchart shows the process used to generate the synthetic clinical notes for our experiments. The source data consisted of open-access, full-text rare disease (RD) case reports available on PubMed. From each RD case report, we manually selected case presentation excerpts that did not reveal the final diagnosis of the RD. The excerpts were then used to prompt the public version of *Gemini 2.5 Flash*, which transformed the excerpts into SOAP-format clinical notes (i.e., with the Subjective, Objective, Assessment, Plan sections). The Subjective and Objective sections from the LLM-generated synthetic notes were extracted and used for testing our homomorphic encryption (HE)-enhanced diagnostic retrieval framework.

### 2.4. Encrypted retrieval

Our retrieval framework integrates functionality from two libraries – *Sentence-Transformer* [10] for transforming text to the embedding vector space and *TenSEAL* [12] for HE, decryption, and mathematic operations in the HE space. From the *Sentence-Transformer* library, our framework inherits the flexibility to choose and deploy from a wide range of embedding LLMs. To generate embedding vectors, free text is chunked to fit within the maximum input sequence limits of the LLM used. Wherever possible, the chunking is done with retaining complete sentences. The contiguous chunks have an adjustable overlap. For retrieval involving multiple input chunks, we used the cosine similarity metric 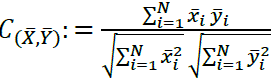; where 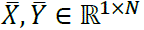 are the average pooled embedding vectors of dimension *N*, with the averaging over *K* vectors derived from *K* chunks is given by 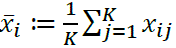 [13,14].

For encryption, we used the Cheon-Kim-Kim-Song (CKKS) HE [15] scheme implemented in the *TenSEAL* library. The configuration used for the CKKS HE scheme are as follows: the *polynomial modulus degree* was 2^13^, which determines the ciphertext capacity and thus controls the security level; the c*oefficient modulus chain* of [60, 40, 40, 60] *bits* was selected to support the required multiplicate depth while preserving numerical precision; a scaling factor of 2^40^was used to enable high-precision during encrypted arithmetic. Galois keys were generated to enable vector products in the HE space. The final retrieval rankings were obtained by decrypting the cosine similarity values computed in the CKKS HE space.

### 2.5. Evaluation

#### Privacy-preserving against inversion attack

For verifying that HE can indeed protect private information compared to raw embeddings, we demonstrated information recovery from unencrypted embedding vectors using the vec2text library [16]. The gtr-t5-base model was first used to generate embedding vectors from excerpts of synthetic clinical notes with the simulated PHI. Subsequently, we used the vec2text’s gtr-t5-base invert_embeddings model to invert the embedding vectors back to the original text. For comparison, we next evaluated whether the same vulnerability exists after encryption, by performing the same inversion attack on the embedding vector that had been encrypted by the CKKS HE scheme.

#### Accuracy of Diagnosis Retrieval

For quantitative evaluation of retrieval performance, the normalized discounted cumulative gain (nDGC) was used. The nDCG measures how well a diagnostic system puts the correct diagnosis near the top of the retrieved list of diseases. The nDCG can take on values between 0 and 1, where a score of 1 indicates the correct diagnosis is ranked first place in the system’s retrieved disease list, and a score of 0 means the retrieved disease list does not contain the correct diagnosis. The exact definition of nDCG is given by: 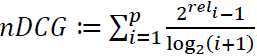; where *rel*_*i*_ ∈ {0,1} is the binary relevance of documents and *p* the position up to which the metric is computed [17]. As a concrete example, say the system retrieved for a test case of known trigeminal neuralgia the ranked disease list: 1) oromandibular dystonia, 2) trigeminal neuralgia, and 3) glossopharyngeal neuralgia; the nDCG for this list would be computed as 0.63, indicating the correct diagnosis is contained in the list but not at the top 1 position.

To assess whether HE alters diagnostic retrieval performance, we performed retrieval on the test data under two conditions: with the query and knowledge base vectors directly without encryption and repeating the same procedure with the vectors encrypted using the CKKS HE scheme. In both settings, the retrieval accuracy was quantified by nDCG on the synthetic test cases. The agreement between the HE and unencrypted retrieval outcomes was compared quantitatively using Kendall’s tau rank correlation coefficient τ [18]. The metric τ quantifies the degree to which the disease rankings with and without encryption agree, by measuring the excess of order-preserved over order-destroyed disease pairs. Kendall’s tau ranges from −1 (representing complete rank reversal), through 0 (representing no rank order association) to +1 (representing identical ordering), with larger absolute values indicating stronger rank order agreement. On top of these, we also compared multiple LLMs (the Embedding Model in **Figure 1**) of varying model sizes and embedding dimensions.

#### Framework robustness under varying parameters

We conducted a series of experiments by varying the embedding and HE parameters to independently assess their impact on the performance of the retrieval framework. Specifically, we varied:

- *Embedding vector dimension*, incremented over 10^1^, 10^2^, 10^3^, and 10^4^. A larger vector dimension implies heavier computing.
- *Polynomial modulus degree*, set to 2^13^, 2^14^, or 2^15^, which represent low, medium, or high security levels respectively. A higher security level increases the dimensionality of the encryption space, making it harder for an adversary to break the encryption computationally.
- *Scaling factor*, set to 2^20^or 2^40^, for low or high precision configurations. Higher precision configurations allow for more significant digits of the data to be preserved during encryption, which would retain the finer semantic nuances in the original embeddings.

For each dimension scale, 100 pairs of embedding vectors were created using a random number generator. Subsequently, for each pair of embedding vectors, cosine similarities were computed under a distinct combination of the *polynomial modulus degree* and *scaling factor* values. The first performance benchmark was *percent deviation d*, defined by:

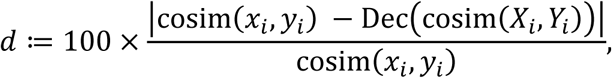

where, the cosine similarity is given by 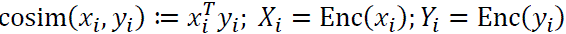. Here Enc(⋅) and Dec(⋅) denote the CKKS encryption and decryption operations respectively. For ease of computations, we use normalized embeddings, *x*_*i*_ and *y*_*i*_, therefore ‖*x*_*i*_‖ = 1 and ‖*y*_*i*_‖ = 1. The other performance benchmark was *time*, measuring the total time needed to encrypt a vector pair, perform cosine similarity in the HE space, and decrypt the result.

## 3. Results

### 3.1. HE blocks the recovery of private information from embeddings

Despite the seeming obfuscation of information offered by projecting text into LLM embedding vectors, we demonstrated that sensitive information can be revealed from unencrypted vectors. As shown in **Table 1**, applying an inversion model to unencrypted embeddings consistently recovered portions of the original text, including the simulated PHI. In contrast, applying the same inversion model on the encrypted embeddings yielded no meaningful or identifiable text. For example, the synthetic patient name “Arthur Miller” in the first row was recovered from the raw vector by the inversion attack, but it was prevented after the vector first went through the HE encryption.

### 3.2. Diagnostic retrieval works in the HE space without performance degradation

The effect of HE over retrieval accuracy is shown in **Figure 3**, where the nDCG distributions with the Qwen3-8B embedding model are plotted across the five RDs. While some RDs are more challenging (e.g., lower nDCGs in ALS and IPF) to the diagnostic retrieval approach, applying HE had essentially negligible effect on the retrieval accuracy within each RD (each pair of the blue versus orange boxplots). The overlap in nDCG measurements along with rank order correlations quantified by Kendall’s tau (τ) consistently equal to 1, indicating that the CKKS HE scheme preserved diagnostic retrieval decisions in the proposed framework.

**Figure 3:**
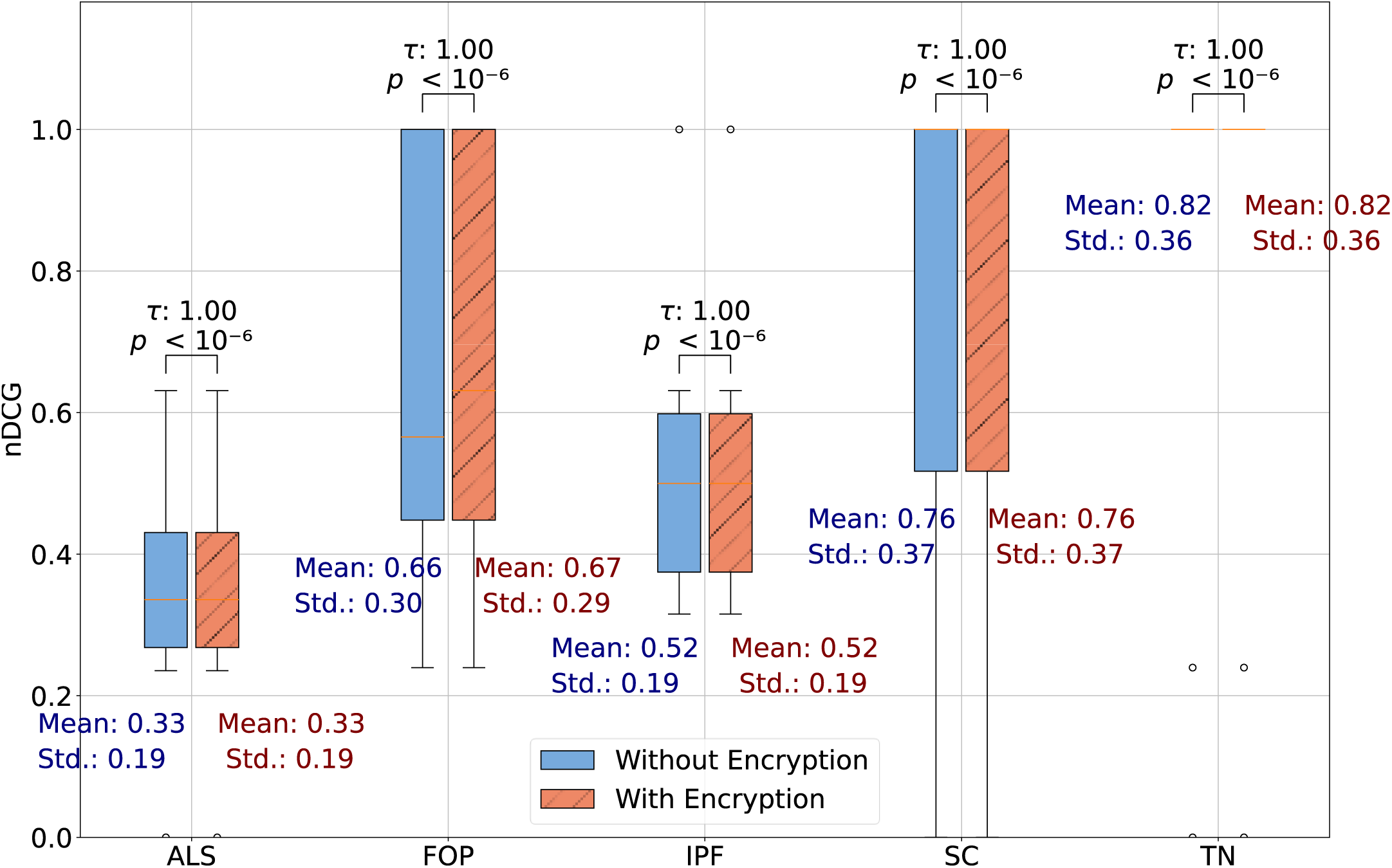
Retrieval performance preserved with homomorphic encryption (HE). Comparative retrieval performances (embedding model: Qwen3-8B) as measured by nDCG are shown with and without HE for the five rare diseases; an nDCG value of 1 indicates perfect retrieval. The lower and upper edges of the box plots represent the 25^th^ and 75^th^ percentiles respectively, the center line the median, and the whiskers the 5^th^ and 95^th^ percentiles. Here, τ is Kendall’s tau rank correlation coefficient. The results indicate that the CKKS HE scheme preserves the retrieval performance. Disease abbreviations: ALS – amyotrophic lateral sclerosis, FOP – fibrodysplasia ossificans progressiva, IPF – idiopathic pulmonary fibrosis, SC – sclerosing cholangitis, TN – trigeminal neuralgia.

### 3.3. Preservation of performance in the HE space is independent from the LLM

To test generalizability beyond a single LLM, we repeated the experiments with and without encryption across four other embedding models that represent a wide range of sizes, complexities, input and embedding dimensions (see details of the LLMs in **SM Table 1**). The consistency in the closely mirroring distributions of nDCG for each model, with τ consistently equal to 1.00 in **Figure 4**, confirms that across all the tested LLMs, encryption left the retrieval performance almost unaltered. This consistency suggests that HE depends more on the embeddings themselves but not the upstream LLM, so the proposed framework can accommodate evolving LLM architectures and perform as good as the raw embeddings’ quality can be. For example, when the raw embeddings yield a leap of nDCG from MiniLM to Qwen3-8B (blue boxplots), that improvement is faithfully reflected in the nDCG by HE-protected embeddings (orange boxplots) with little loss.

**Figure 4:**
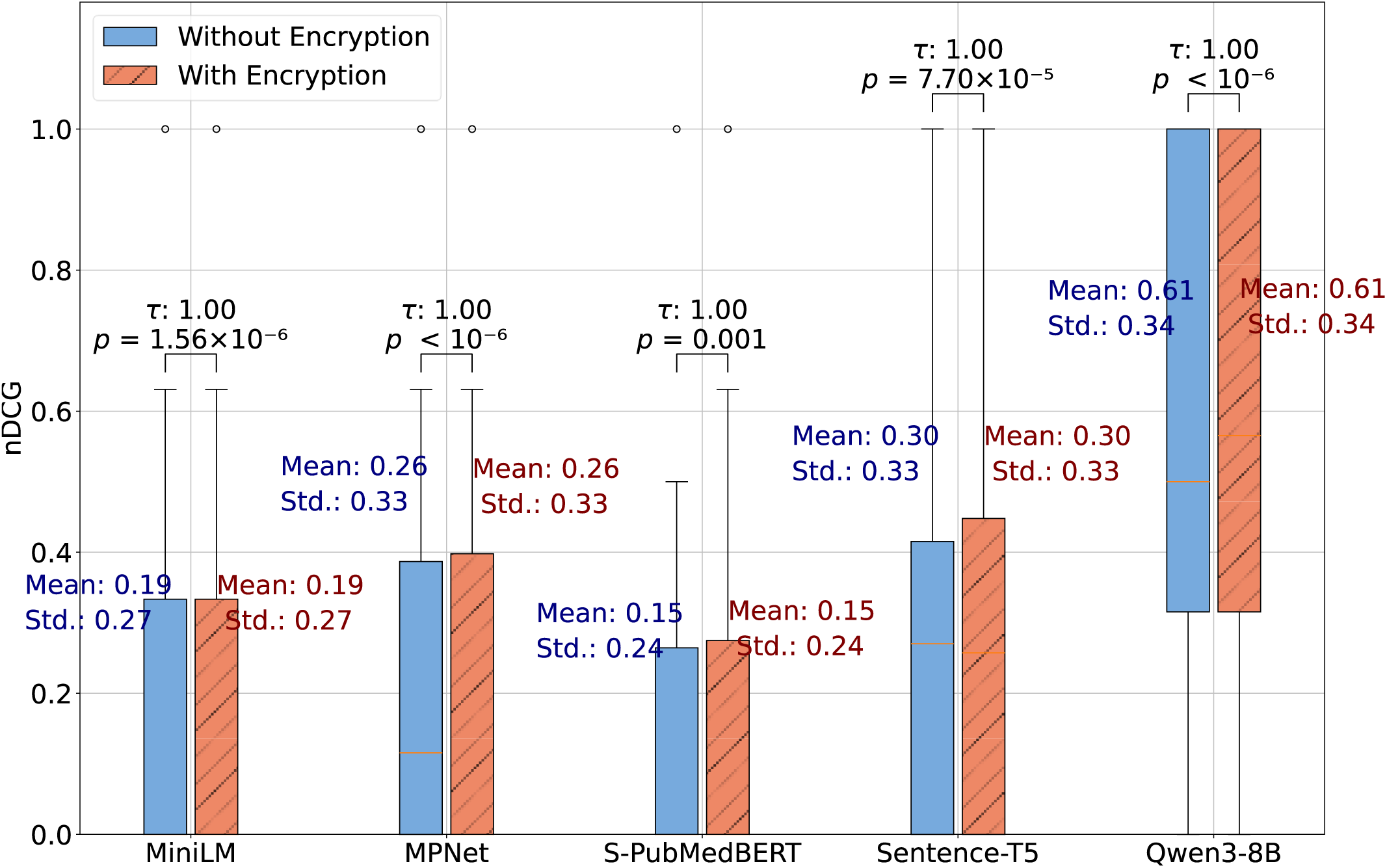
Preservation of retrieval performance under homomorphic encryption (HE) does not rely on a specific large language model (LLM). For each LLM, the retrieval performance is based on pooling the results across all five tested rare diseases. Here, τ is Kendall’s tau rank correlation coefficient that measures the diagnostic agreement between the HE and no-HE results. The complexity of the LLMs roughly increment from left to right, e.g., the embedding dimensions range from 384 for the MiniLM model to 4096 for the Qwen3-8B model. Regardless of the LLM used, consistently the HE scheme did not hinder the retrieval performance compared to no encryption.

### 3.4. The deviation due to encryption can be tuned down with modest computation cost

The effects of the CKKS HE parameters on numerical deviations and computation costs are illustrated in **Figure 5**. The trends in **Figure 5 A** show that higher security settings increased the deviation introduced by HE on cosine similarity computations by approximately a factor of 10, compared to lower security settings at a fixed precision level. In contrast, increasing the precision, substantially reduced the deviation by a factor of 1000. Specifically, the high precision plus high security combined settings resulted in deviations less than 10 parts per million for all vector sizes. In **Figure 5 B,** it is observed that the computation times, even for the most expensive settings (high-security and high-precision), consistently remain below 0.5s per cosine similarity computation even for the largest vector pairs tested (a dimension of 10,000). These findings provide evidence that our proposed framework can accommodate a balance in security and accuracy without compromising implementation feasibility.

**Figure 5:**
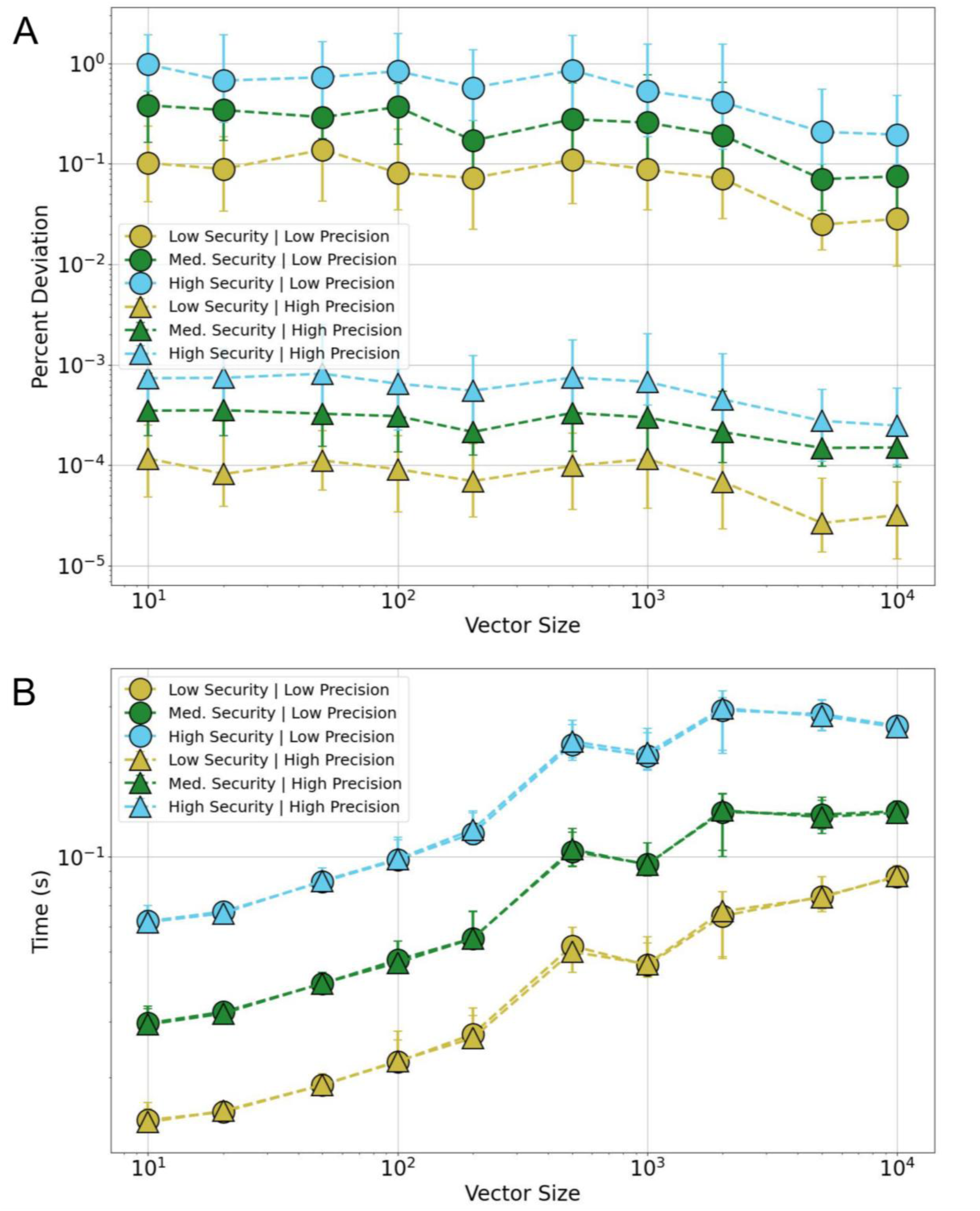
Balancing accuracy, security, and computation time under homomorphic encryption (HE). The effect of the encryption settings (three Security levels and two Precision levels) for similarity-based retrieval is presented. Each data marker represents the median cosine similarity computed across 100 independent simulation runs, wherein, each run used a new and distinct encryption key. The whiskers mark the 25^th^ and 75^th^ percentiles. The Y-axis stands for a different measure in the individual experiment: (A) The deviations introduced by encryption as a function of vector size and CKKS scheme parameters; (B) Computation time per encrypted cosine similarity computation that corresponds to varying vector size and CKKS scheme parameters. Note: both the vertical as well the horizontal axes are shown in logarithmic scale.

## 4. Discussion

We developed a privacy-preserving information retrieval framework integrating HE and LLMs to differentially rank the top possible diagnoses by screening through a comprehensive knowledge base with thousands of RDs. Our experiments confirmed that HE protects against inversion attacks [15, 21] without compromising the rich information in embeddings. The framework works for any LLM as the embedding model while offering tunable parameters to balance the deviation due to encryption, security strength, and computational efficiency. The efficiency can be further boosted by parallelizing independent operations such as the similarity computations. Collectively, these merits make it a practical privacy-preserving layer for AI-assisted decision-support solutions. Moreover, the client-server architecture is versatile and can be easily generalized to other applications such as clinical trial criteria matching. In the translational science context, mature implementation of such a framework would encourage serving trustable AI solutions at resource-rich hubs, which could be institutions or federal agencies specialized in a disease area. There is economic benefit in mitigating redundant AI hosting, and the associated reduction of carbon footprint follows as an environmental benefit.

Compared with conventional privacy-enhancing techniques, the proposed framework offers several advantages. First, it eliminates the need for deidentification, a process that remains resource-intensive and technically challenging [20]. In data-rich and networked environments, even datasets deidentified under the HIPAA Safe Harbor standard have been shown to be re-identifiable, indicating the defects of current approaches and the likelihood of increasingly stringent regulation in the future [21]. Second, our approach alleviates the financial burden of performing deidentification prior to using external AI and/or hosting expensive AI services on premises, thus freeing up budget for alternative investments. Third, the encrypted retrieval process preserves the raw medically relevant information in its intact form, maintaining diagnostic interpretability (matched information can be decrypted by the user or an authorized party) while safeguarding patient privacy [6].

We identify the following limitations in the pilot study: 1) The CKKS encryption scheme does not preserve ranking order and leaves that to client-side postprocessing after decryption. However, such computational overhead is modest and can be further optimized. 2) The study only used synthetic clinical notes of a few diseases to prove the technical feasibility of integrating HE and LLMs; future work is needed to evaluate the framework on real patient data and assess the robustness subject to broader medical conditions and noise such as typos and negations. 3) The methodology is demonstrated only for text-based data, and extension to other data modalities, such as images and signals remains to be explored. 4) We did not evaluate the full spectrum of potential threats against a system secured by HE, such as theft of the key, side-channel and fault-injection attacks [22]. A dedicated security-assessment study is required before deployment in real-world clinical environments.

## 5. Conclusions

We demonstrate a privacy-preserving retrieval framework that integrates HE with LLMs, using RD diagnosis as an example task. The HE encryption can protect privacy without hindering diagnostic retrieval performance, exemplifying an overlooked yet effective privacy-preserving technique that can benefit the modern AI implementation stack. Our framework should facilitate the translation of healthcare AI by simultaneously incentivizing centralization (of high-quality services) and democratization (of distributed direct accesses) through effective and affordable privacy protection.

## Contributions

**Sivaraman Rajaganapathy:** Methodology, Experiment design and execution, Writing.

**Jennifer St. Sauver:** Writing – critical review and editing. **Filippo Pinto e Vairo:** Writing – critical review and editing. **Prasad G. Iyer:** Witing – critical review and editing. **Hongfang Liu:** Writing – critical review and editing. **Jungwei W. Fan:** Conceptualization, Methodology, Experiment design, Resources, Supervision, Writing.

## Data and Code Availability

The source codes, instructions, and data presented in this manuscript are available upon request.

## Data Permissions

The Orphadata Science dataset was used in this study in accordance with the terms stipulated by the CC BY 4.0 licence.

## Data Availability

All data produced in the present study are available upon reasonable request to the authors.

## Supplementary Material

**SM Table 1:** A summary of the LLMs experimented for creating the embedding vectors.

| **Model** | **Maximum**<br>**Input**<br>**Sequence**<br>**Length**<br>**(Tokens)** | **Dimension of**<br>**Embedding**<br>**Vector** | **Size of**<br>**Model**<br>**(MB)** | **Additional Details** |
| --- | --- | --- | --- | --- |
| MiniLM<br>(all MiniLM-L6-V2) | 256 | 384 | 80 | General purpose<br>embedding model trained<br>on more than 1 billion<br>training pairs. |
| MPNet<br>(all-mpnet-base-v2) | 384 | 768 | 420 | General purpose<br>embedding model trained<br>on more than 1 billion<br>training pairs. |
| S-PubMedBERT<br>(S-PubMedBERT-MS-MARCO) | 350 | 768 | 438 | BiomedNLP<br>PubMedBERT model fine-<br>tuned on MS-MARCO<br>dataset. |
| Sentence-T5<br>(Sentence-t5-xxl) | 256 | 768 | 9,230 | T5-11B model trained on<br>2 billion sentence<br>similarity tasks. |
| Qwen3-8B<br>(Qwen3-Embedding-8B) | 32,000 | 4,096 | 15,130 | Proprietary model of the<br>Qwen family. |

